# Impaired performance of SARS-CoV-2 antigen-detecting rapid tests at elevated temperatures

**DOI:** 10.1101/2021.01.06.21249314

**Authors:** Verena Haage, Edmilson Ferreira de Oliveira-Filho, Andres Moreira-Soto, Arne Kühne, Carlo Fischer, Jilian Sacks, Victor Max Corman, Marcel A. Müller, Christian Drosten, Jan Felix Drexler

**Author notes:** Correspondence: Jan Felix Drexler, Helmut-Ruska-Haus, Institute of Virology, Campus Charité Mitte, Charitéplatz 1, 10098 Berlin, Germany.

## Abstract

Rapid antigen-detecting tests (Ag-RDTs) can complement molecular diagnostics for COVID-19. The recommended temperature for storage of SARS-CoV-2 Ag-RDTs ranges between 5-30°C. In many countries that would benefit from SARS-CoV-2 Ag-RDTs, mean temperatures exceed 30°C. We assessed analytical sensitivity and specificity of eleven commercially available SARS-CoV-2 Ag-RDTs using different storage and operational temperatures, including (i) long-term storage and testing at recommended conditions, (ii) recommended storage conditions followed by 10 minutes exposure to 37°C and testing at 37°C and (iii) 3 weeks storage followed by testing at 37°C. The limits of detection of SARS-CoV-2 Ag-RDTs under recommended conditions ranged from 8.2×10^5^-7.9×10^7^ genome copies/ml of infectious SARS-CoV-2 cell culture supernatant. Despite long-term storage at recommended conditions, 10 minutes pre-incubation of Ag-RDTs and testing at 37°C resulted in about ten-fold reduced sensitivity for 46% of SARS-CoV-2 Ag-RDTs, including both Ag-RDTs currently listed for emergency use by the World Health Organization. After 3 weeks of storage at 37°C, 73% of SARS-CoV-2 Ag-RDTs exhibited about ten-fold reduced sensitivity. Specificity of SARS-CoV-2 Ag-RDTs using cell culture-derived human coronaviruses HCoV-229E and HCoV-OC43 was not affected by storage and testing at 37°C. In summary, short- and long-term exposure to elevated temperatures likely impairs sensitivity of several SARS-CoV-2 Ag-RDTs that may translate to false-negative test results at clinically relevant virus concentrations compatible with inter-individual transmission. Ensuring appropriate transport and storage conditions, and development of tests that are more robust across temperature fluctuations will be important for accurate use of SARS-CoV-2 Ag-RDTs in tropical settings.

## Introduction

Advantages of SARS-CoV-2 antigen-detecting rapid diagnostic tests (Ag-RDTs) include fast results and their applicability on site without dependence on laboratory settings. Ag-RDTs are not intended to replace real-time RT-PCR as the gold standard for SARS-CoV-2 diagnostics (1, 2), but can be deployed for surveillance in high-risk environments or areas with insufficient laboratory capacity. With a constantly growing number of commercially available Ag-RDTs on the global market, the number of studies validating Ag-RDTs from different manufacturers is increasing rapidly (3-9). Those validation studies have so far not focused on the performance of Ag-RDTs under conditions that differ from recommended storage and test execution conditions (5-30°C), such as those observed in tropical settings where ambient temperatures commonly exceed 30°C (**Figure 1A**). This is challenging because tropical regions are strongly affected by the SARS-CoV-2 pandemic as evident from total cases reported from India, Brazil, Argentina, and Colombia, four out of the ten most affected countries worldwide by November 2020 (**Figure 1B**).

**Figure 1.**
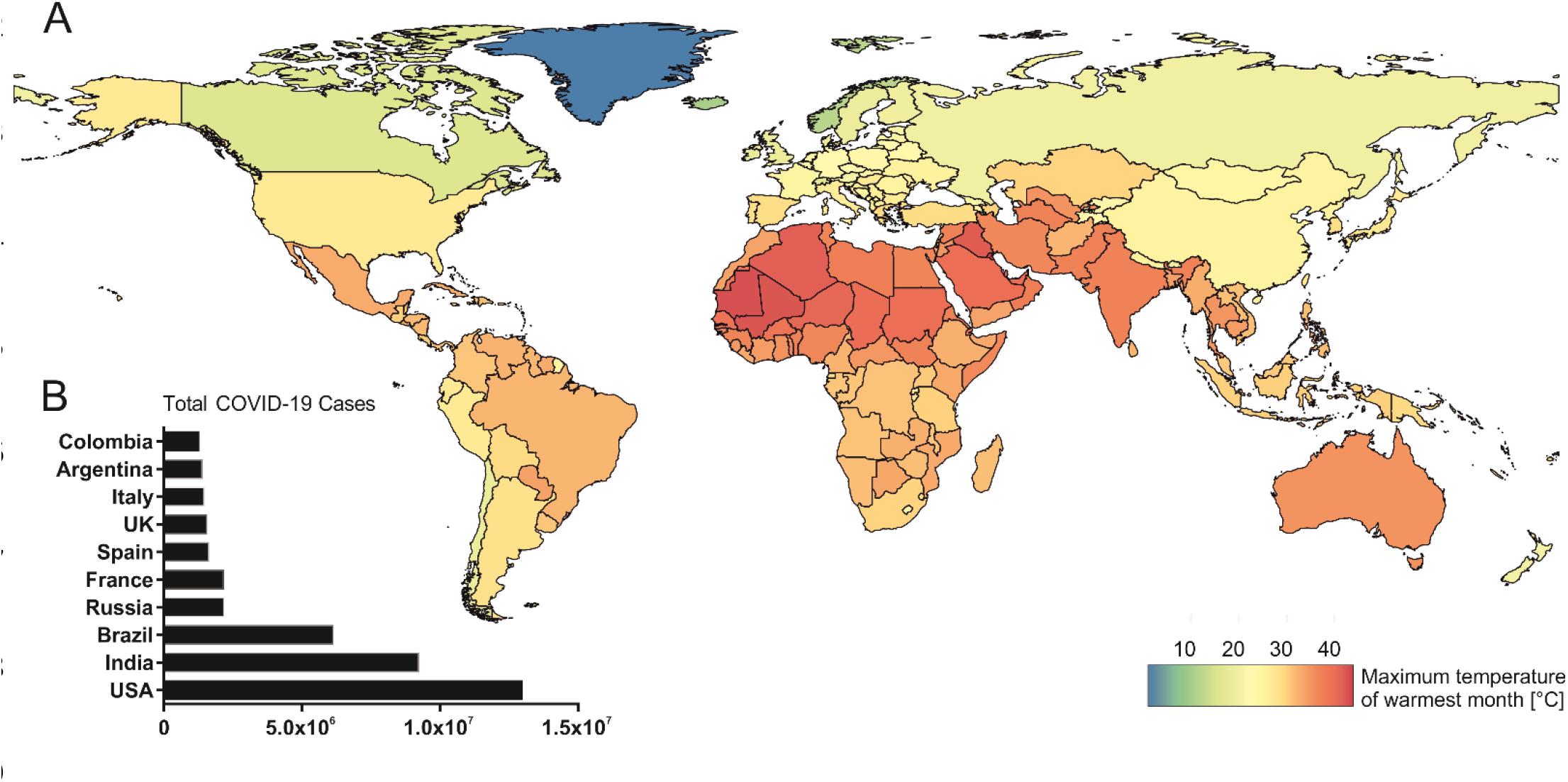
COVID-19 case numbers and mean temperatures globally. A. World map representing global temperature distribution based on maximum temperature of the warmest month (°C) freely available from WorldClim 2 (19). B. Graph represents total COVID-19 cases in the ten most affected countries globally by 25^th^ of November 2020 (20).

Several studies have shown the need for validation of diagnostic tools in resource-limited tropical regions because co-endemic diseases can affect diagnostic test performance (10-14). The same might also apply to the environmental conditions under which Ag-RDTs are transported, stored and used. Temperature tolerance of SARS-CoV-2 diagnostic tools or environmental stability requirements have been previously discussed as hurdles and corresponding guidelines for stability testing of *in vitro* diagnostics are provided by the World Health Organization (WHO) (15, 16). To date, knowledge about temperature stability of SARS-CoV-2 Ag-RDTs is scarce.

To validate the performance of SARS-CoV-2 Ag-RDT in tropical settings, we compared analytical sensitivity and specificity of eleven SARS-CoV-2 Ag-RDTs using recommended and elevated temperatures.

## Materials and Methods

### Storage and testing of SARS-CoV-2 Ag-RDTs at 37°C

For storage at recommended conditions, the tests were stored at ambient temperature and were monitored that the room temperature stayed between 15-25°C. For storage at 37°C, SARS-CoV-2 Ag-RDTs were removed from cartons for space reasons and stored in plastic bags in a 37°C incubator for 19-21 days for sensitivity and 8-9 days for specificity testing. All tests were kept in their individual foil packaging until use. For experiments with infectious SARS-CoV-2, Ag-RDTs were transferred to a BSL-3 laboratory and stored in a 37°C incubator until test application. For specificity testing with infectious human coronavirus (HCoV)-229E and HCoV-OC43, SARS-CoV-2 Ag-RDTs were transferred to a BSL-2 laboratory and stored in a 37°C incubator until test validation. For experiments testing short-term exposure to 37°C following storage at recommended temperatures, SARS-CoV-2 Ag-RDTs were transferred to a BSL-3 laboratory and stored for 10 minutes in a 37°C incubator until test operation. After application of SARS-CoV-2, HCoV-229E or HCoV-OC43 cell culture supernatant in a biological safety cabinet, Ag-RDTs were further incubated in a 37°C incubator until readout of the results.

### Analytical sensitivity

SARS-CoV-2 (BetaCoV/Munich/ChVir984/2020) was grown on Vero E6 cells (C1008; African green monkey kidney cells) and maintained in DMEM containing 10% FCS at 37°C with 5% CO2. Viral RNA was extracted from cell culture supernatant using the QIAamp Viral RNA Mini Kit (Qiagen, Hilden, Germany) according to the manufacturer’s instructions. RNA concentration was determined by specific real-time RT-PCR and *in vitro*-transcribed RNA standards designed for absolute quantification of virus concentrations (17, 18).

For determination of the limit of detection (LOD), a SARS-CoV-2 stock (3.1×10^9^ copies/ml) was serially diluted in plain DMEM and 5µl/dilution were added to the extraction buffer of the respective kit for validation experiments. For the Coris COVID-19 Ag Respi-Strip test, 5µl of SARS-CoV-2 supernatant was added to 95µl of PBS to reach the required sample volume of 100µl prior to addition of LY-S buffer to the sample for test validation. Initially, validation experiments were performed in triplicates for a small subset of tests (ActivXpress, Genedia, ichroma, JOYSBIO) at recommended conditions (i) with all three replicates showing the same result (**Supplementary Table 1**). Consequently, validation experiments for all included tests were subsequently performed in duplicates due to a limited amount of available tests for all conditions tested. LOD was defined as the lowest dilution at which both replicates were positive. A dilution factor correction was applied based on the volume of extraction buffer (range between kits, 100-500µl) provided by each SARS-CoV-2 Ag-RDT kit.

### Analytical specificity

Specificity was tested using cell culture supernatant of HCoV-229E and HCoV-OC43, grown on CaCo-2 cells. Quantification of viral stocks was performed by RT-PCR using an in house protocol and photometrically quantified *in vitro*-transcribed RNA standards as described above. 5µl of stocks were directly used for validation experiments. The required sample volume of 100µl for the Coris COVID-19 Ag Respi-Strip was reached as described above. Experiments were performed in duplicates.

### Interpretation of test results

Tests were performed according to the test-specific supplier instructions for use, except for the sample preparation as noted above. Results in the form of a band on immunochromatography paper were scored independently by two persons except for the reader-based tests Bioeasy 2019-nCoV Ag and ichroma - COVID-19 Ag. In case of discrepancy for visual tests, a third person was consulted to reach a final decision. Results were defined as unclear when a weak, discontinuous band or smear was observed that could not be clearly defined as a positive or negative result. In case of test failure indicated by absence of a visible positive control band, the test procedure was repeated with the same sample.

### World heat map

Data of maximum temperatures of the hottest month (°C) on country level at the spatial resolution of 2.5 min were obtained from WorldClim 2 (19). National means were calculated using the exactextractr package in R version 4.0.2. Data on COVID-19 cases were obtained from Worldmeter (20) and visualized using the GraphPad Prism software.

## Results

At present, there are at least 119 SARS-CoV-2 Ag-RDTs commercially available (21). Eleven of those SARS-CoV-2 rapid antigen tests were selected for temperature stability validation based on the availability of clinical performance data (3), and manufacturing by leading suppliers implying availability on the global market (**Table 1**). Subsequently, analytical performance of selected SARS-CoV-2 Ag-RDTs was assessed following storage and application of tests under recommended conditions as well as elevated temperatures (termed tropical conditions henceforth), using three different experimental settings.

**Table 1.** Overview of SARS-CoV-2 rapid antigen tests included in the study.

| ID | Test | Manufacturer | Readout | Lot No. |
| --- | --- | --- | --- | --- |
| I | Panbio™ COVID-19 Ag Rapid Test | Abbott Laboratories | Visual | 41ADF012A |
| II | ActivXpress + COVID-19 Antigen Complete Testing Kit | Edinburgh Genetics | Visual | AG20200905 |
| III | Bioeasy 2019-nCoV Ag Fluorescence Rapid Test Kit | Shenzhen Bioeasy Biotechnology Co., Ltd | Reader | 2003N406 |
| IV | Clinitest Rapid COVID-19 Antigen Test | Siemens Healthineers | Visual | 2010184 |
| **V** | Covid.19 Ag Respi-Strip | Coris BioConcept | Visual | 43871J2008<br>43760I2015 |
| **VI** | COVID-19 Ag | Genedia | Visual | 643X2005 |
| **VII** | ichroma - COVID-19 Ag | Boditech Med | Reader | SRQHA27 |
| **VIII** | COVID-19 Antigen Rapid Test Kit | JOYSBIO (Tianjin) Biotechnology Co., Ltd. | Visual | 2020092409 |
| **IX** | NowCheck COVID-19 Ag test | BIONOTE INC. | Visual | 1901D002 Code GEN |
| **X** | SARS-CoV-2 Rapid Antigen Test | Roche Diagnostics | Visual | QCO3020083<br>QCO390003I/Sub:I-2<br>QCO390011A/Sub:A-2 |
| **XI** | STANDARD Q COVID-19 Test | SD Biosensor, Inc. | Visual | QCO3020040A |

The first setting was (i) storage at recommended conditions (room temperature (rt); 15-30°C) for 1-6 months and test incubation at recommended conditions (rt; 15-30°C). The second setting was (ii) storage at recommended conditions (rt; 15-30°C) for 1-6 months, 10 minutes pre-incubation of tests at 37°C prior to testing and test incubation at 37°C so as to mimic recommended storage of kits prior to test usage under non air-conditioned conditions in tropical settings. The third setting was (iii) storage under tropical conditions (37°C) for 19-21 days followed by test incubation at 37°C to mimic non air-conditioned storage and test application in tropical settings (**Figure 2**).

**Figure 2.**
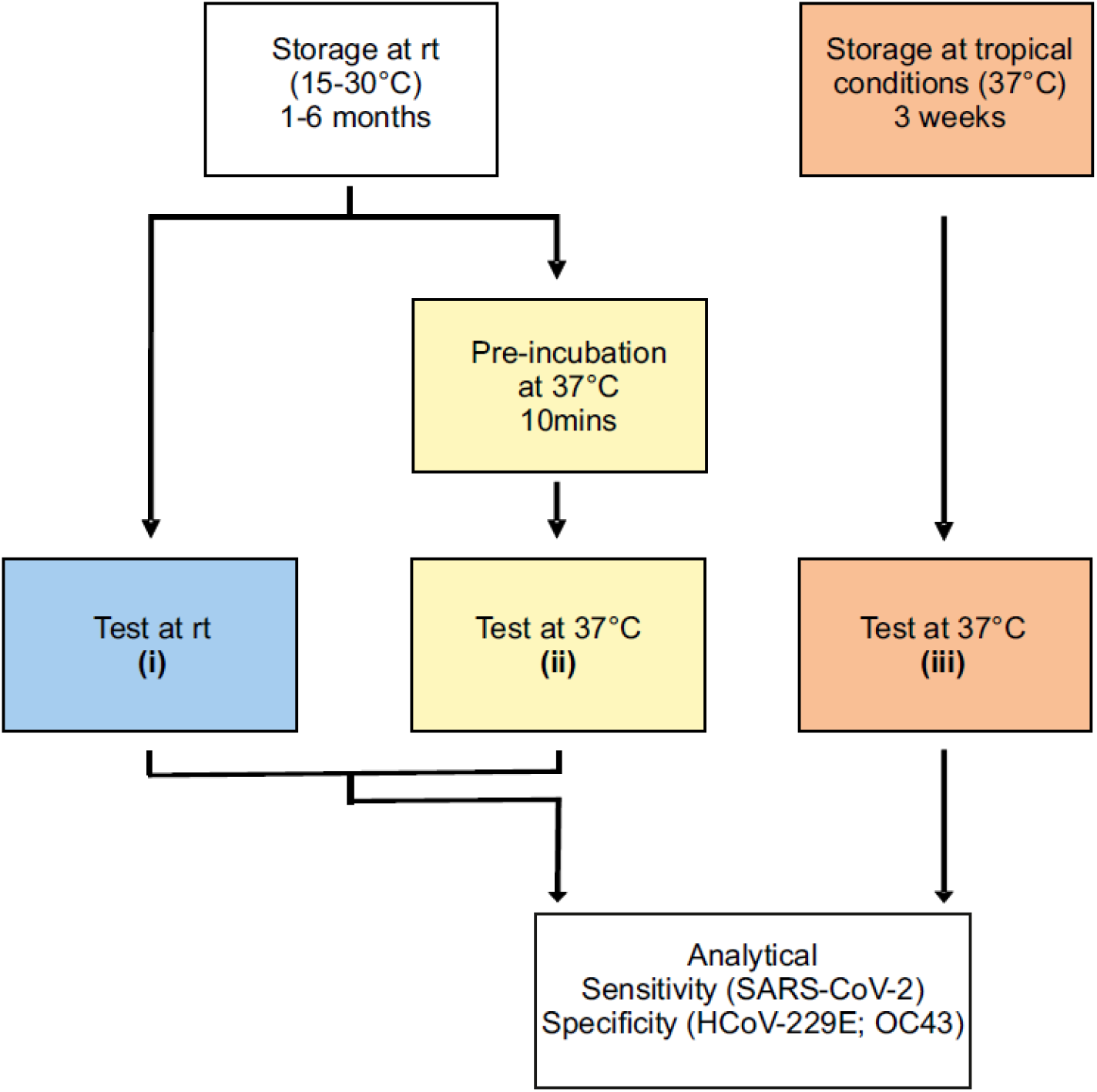
Experimental setup for validation of SARS-CoV-2 Ag-RDTs. (i): storage and operation at recommended conditions; (ii): recommended storage and operation at tropical conditions; (iii): storage and operation at tropical conditions.

First, we determined analytical sensitivity at recommended conditions by determining the limit of detection (LOD) of SARS-CoV-2 Ag-RDTs at room temperature using serial dilutions of SARS-CoV-2 cell culture supernatant (dilution range: 7.2×10^6^ copies/ml to 3.1×10^9^ copies/ml; **Table 2**) following storage for 1-6 months at room temperature. Prior to performing all experiments in duplicates we validated analytical sensitivity for a small subset of tests at recommended conditions in triplicates, showing that results for duplicates and triplicates were identical (**Supplementary Table 1**). As extraction buffer volumes differ between manufacturers and a constant volume of SARS-CoV-2 supernatant was applied for validation experiments, the limit of detection (LOD) for each Ag-RDT was calculated by correcting for the respective dilution factor. The dilution-factor corrected limit of detection (LOD) for validated SARS-CoV-2 Ag-RDTs ranged from 8.2×10^5^ copies/ml to 7.9×10^7^ copies/ml of SARS-CoV-2 cell culture supernatant (**Table 2**). Those LODs were consistent with previously published virus concentrations for validation of Ag-RDTs using clinical samples (3), suggesting robustness of our data. Our data also highlight profound differences in analytical sensitivity of up to 100-fold for SARS-CoV-2 Ag-RDTs from different manufacturers.

**Table 2.**
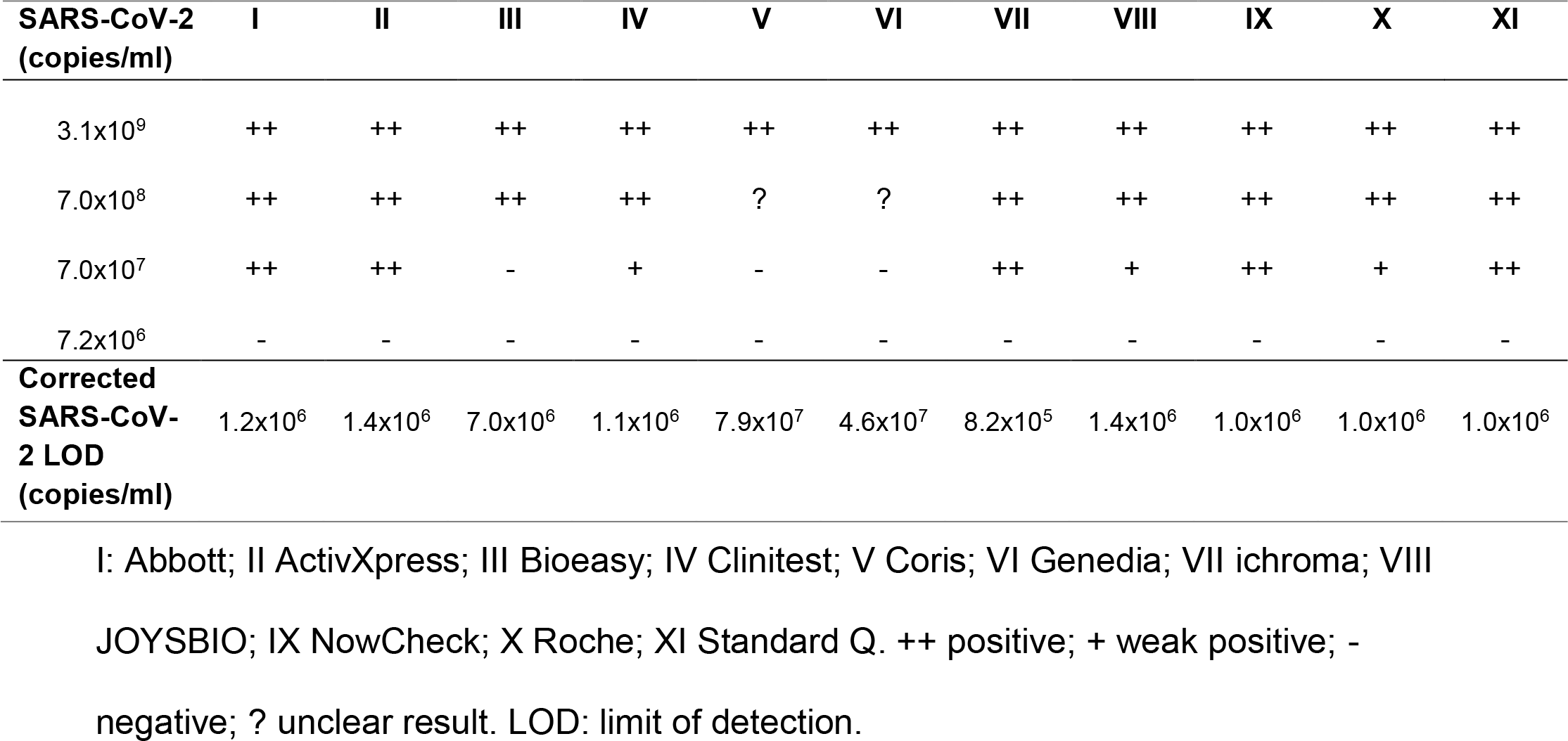
Analytical Sensitivity of SARS-CoV-2 rapid antigen tests at recommended storage and test conditions.

We then assessed analytical sensitivity of SARS-CoV-2 Ag-RDTs following short- and long-term exposure to 37°C (settings (ii) and (iii); **Figure 2**) by determining the level of detection (LOD) using identical serial dilutions of SARS-CoV-2 cell culture supernatant. The analytical sensitivity of about half of the evaluated SARS-CoV-2 Ag-RDTs (five out of eleven; 46%) was already compromised by about ten-fold when tests were stored under recommended conditions but exposed to 37°C for only ten minutes prior to testing at 37°C (**Figure 3**). After 19-21 days storage at 37°C and testing at 37°C, eight out of the eleven SARS-CoV-2 Ag-RDTs (73%) showed an about ten-fold reduction in analytical sensitivity when compared to recommended temperatures.

**Figure 3.**
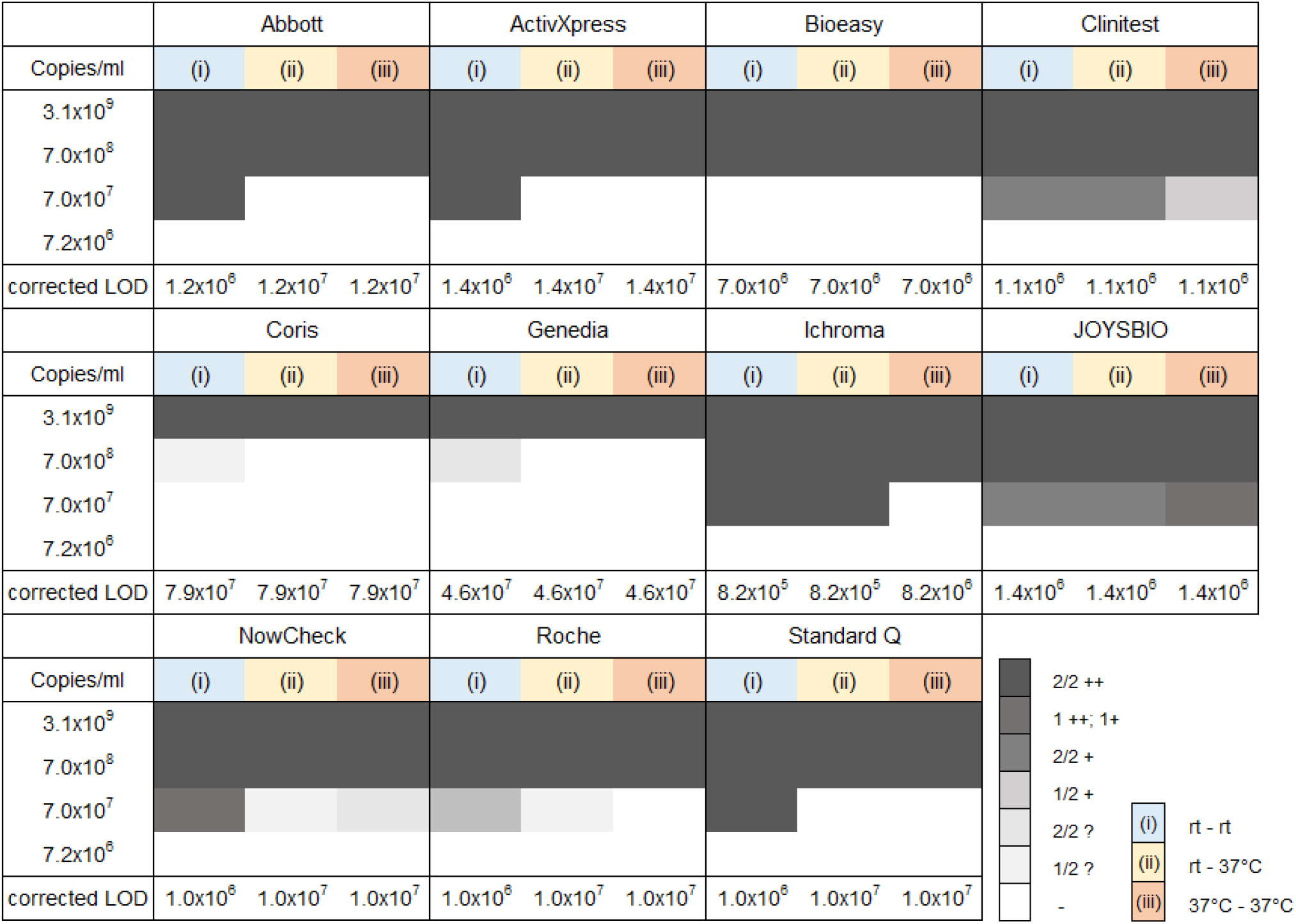
Sensitivity of SARS-CoV-2 Ag-RDTs decreases at elevated temperatures. Analytical sensitivity of SARS-CoV-2 rapid antigen tests upon different storage and operation conditions; (i): rt storage-rt testing, blue; (ii): rt storage - 37°C 10 min pre-incubation - 37°C testing, yellow; (iii): 37°C storage - 37°C testing, orange. ++ positive; + weak positive; - negative; ? unclear result. 1/2, 2/2: number of replicates. LOD: limit of detection. rt: room temperature.

Additionally, analytical specificity of SARS-CoV-2 Ag-RDTs under recommended and under tropical storage and test application conditions ((i) and (iii); **Figure 2**) was examined by testing for cross-reactivity with the ubiquitous HCoV-229E (2.9×10^7^ copies/ml) and HCoV-OC43 (1.0×10^6^ copies/ml) (22, 23). Those HCoV concentrations were selected according to the guidelines on analytical specificity testing for SARS-CoV-2 Ag-RDTs published by the German Federal institute for vaccines and biomedicines (24). SARS-CoV-2 Ag-RDTs showed no cross-reactivity with HCoV-229E or HCoV-OC43 upon storage and testing at elevated temperatures (**Table 3**).

**Table 3.** **Analytical Specificity of SARS-CoV-2 rapid antigen tests at recommended and tropical storage and test application conditions**

| Condition | Virus | Cps/ml | I | II | III | IV | V | VI | VII | VIII | IX | X | XI |
| --- | --- | --- | --- | --- | --- | --- | --- | --- | --- | --- | --- | --- | --- |
| (i) | HCoV-229E | $2.9 \times 10^7$ | - | - | - | - | - | - | - | - | - | - | - |
| (iii) |  |  | - | - | - | - | - | - | - | - | - | - | - |
| (i) | HCoV-OC43 | $1.0 \times 10^6$ | - | - | - | - | - | - | - | - | - | - | - |
| (iii) |  |  | - | - | - | - | - | - | - | - | - | - | - |

I: Abbott; II ActivXpress; III Bioeasy; IV Clinitest; V Coris; VI Genedia; VII ichroma; VIII JOYSBIO; IX NowCheck; X Roche; XI Standard Q.+ positive; - negative. Cps, Genome copies.

## Discussion

Our study highlights that even short-term exposure to elevated temperatures may compromise sensitivity of currently available SARS-CoV-2 Ag-RDTs.

Our data are consistent with impaired sensitivity of other Ag-RDTs at elevated temperatures, including Malaria rapid diagnostic tests (MRDTs). An assessment of five MRDTs reported a 13% to 53% decline in sensitivity for three MRDTs following 90 days of storage at 35°C (25). Moreover, an evaluation assessing temperature stability of dengue NS1 antigen-based RDTs at 35°C showed a gradual decline in test sensitivity for seven out of eight tested dengue Ag-RTDs after storage for about 20 days at elevated temperatures (26). Beyond storage, elevated temperatures during shipment can also affect Ag-RDT performance. Supply chains of MRDTs were studied in Burkina Faso, Senegal, Ethiopia, the Philippines and Cambodia, demonstrating regular exceeding of 30°C during transport (27, 28). Consequently, the WHO recommends heat stability testing between 35°C and 40°C for MRDTs (29) and supply and delivery chains to tropical countries must contain adequate cold chains (30).

As discussed by others, temperature stability guidelines for *in vitro* diagnostics exist, however there are currently no specific guidelines for the validation of Ag-RDTs regarding temperature stability (31-34). Common validation guidelines including environmental conditions could be a first step towards globally reliable diagnostics.

At the same time, our data imply a huge challenge to tropical countries with regard to adequate storage of SARS-CoV-2 Ag-RDTs. Besides storage capacity, temperature control and monitoring of storage facilities is required, especially for long-term storage of Ag-RDTs. To guarantee temperature-regulated storage, a certain level of infrastructure is required, ideally air-conditioned facilities with temperature monitors and secured power supply. However, these requirements are not realistic and appropriate concepts for adequate storage in remote areas without electricity and rudimentary infrastructure will be required.

Our study is limited by focusing on analytical test performance exclusively for reasons of comparability of test results across the different conditions and based on limited access to clinical samples. An additional limitation of our study is the use of duplicates for some tests instead of a higher number of replicates, which was due to the limited availability of all tests included in the study. Further studies will be required to assess test performance upon tropical storage and application in tropical conditions using large numbers of clinical samples. Additionally, not only the effect of elevated temperature but also the effect of increased humidity on test performance will have to be assessed. Finally, future studies documenting the impact of decreased temperature on test performance may also be needed. Despite these limitations, our study presents a robust resource for further validation studies as a high number of SARS-CoV-2 Ag-RDTs was included. Additionally, our data on an overall impaired performance of Ag-RDTs at elevated temperatures are consistent across tests and analytical sensitivity for several tests was identical upon usage of either duplicates or higher numbers of replicates.

In sum, it was shown previously that clinically relevant virus concentrations of about 10^6^ genome copies per ml suffice for virus isolation and culture and therefore serve as a correlate for infectivity (35, 36). Our study strongly suggests that short- and long-term exposure to elevated temperatures may compromise sensitivity of SARS-CoV-2 Ag-RDTs to an extent that may lead to false-negative test results at clinically relevant virus concentrations, potentially enhancing SARS-CoV-2 spread in tropical settings.

## Data Availability

All relevant data are within the manuscript and its Supporting Information files.

## Author Contributions

V.H.: conceptualization, investigation, validation, formal analysis, data curation, writing – original draft preparation, visualization. E.F.: methodology, investigation, validation. A.M.S.: investigation, validation, visualization. A.K.: investigation. C.F.: visualization, software. J.S.: methodology, resources, writing. V.M.C.: methodology. C.D.: methodology. J.F.D.: conceptualization, methodology, resources, writing – original draft preparation, visualization, supervision, project administration, funding acquisition. All authors have read and agreed to the published version of the manuscript.

---

**Supplementary Table 1.** Analytical Sensitivity of SARS-CoV-2 rapid antigen tests at recommended storage and test conditions.

| Test | ActivXpress |  |  | Genedia |  |  | ichroma |  |  | JOYSBIO |  |  |
| --- | --- | --- | --- | --- | --- | --- | --- | --- | --- | --- | --- | --- |
| Replicate | 1 | 2 | 3 | 1 | 2 | 3 | 1 | 2 | 3 | 1 | 2 | 3 |
| **SARS-CoV-2**<br>**(copies/ml)** |  |  |  |  |  |  |  |  |  |  |  |  |
| 3.1x10<sup>9</sup> | ++ | ++ | ++ | ++ | ++ | ++ | ++ | ++ | ++ | ++ | ++ | ++ |
| 7.0x10<sup>8</sup> | ++ | ++ | ++ | ? | ? | ? | ++ | ++ | ++ | ++ | ++ | ++ |
| 7.0x10<sup>7</sup> | ++ | ++ | ++ | - | - | - | ++ | ++ | ++ | + | + | + |
| 7.2x10<sup>6</sup> | - | - | - | - | - | - | - | - | - | - | - | - |

++ positive; + weak positive; - negative; ? unclear result. LOD: limit of detection.

